# CBCRisk-Mastectomy: A Risk Prediction Tool to Aid Contralateral Prophylactic Mastectomy Decision Making

**DOI:** 10.64898/2026.05.12.26352924

**Authors:** Ibrahim Hossain Sajal, Ruth M. Pfeiffer, Ismail Jatoi, Mitchell H. Gail, Reena S. Cecchini, Pankaj K. Choudhary, Swati Biswas

## Abstract

**Purpose:** Unilateral breast cancer (BC) patients scheduled for mastectomy often choose to undergo contralateral prophylactic mastectomy (CPM), despite substantial declines in contralateral breast cancer (CBC) risk in recent decades. Models predicting absolute risk of future CBC can aid informed decision-making about CPM. CBCRisk is an existing CBC absolute risk prediction model trained on unilateral BC patients regardless of whether they had mastectomy. Here we developed CBCRisk-Mastectomy, tailored specifically to BC patients scheduled for mastectomy and considering CPM.

**Patients and Methods:** We used data on BC patients who underwent mastectomy to treat their first BC from two nationally representative sources: Breast Cancer Surveillance Consortium (BCSC) and Surveillance, Epidemiology, and End Results (SEER) cancer registry. We imputed missing data in the BCSC sample and used conditional logistic regression models, trained on 2,660 BC patients (665 CBC cases) from BCSC, to identify predictors and estimate relative risks (RRs). These were combined with attributable risks and CBC incidence rates estimated from SEER to obtain absolute risk. Cross-validation was used to internally validate CBCRisk-Mastectomy and compare with CBCRisk.

**Results:** CBCRisk-Mastectomy has nine predictors: first BC type, lobular carcinoma in situ status, estrogen receptor status, tumor stage, breast density, age at BC diagnosis, family history of BC, age at first birth, and body mass index. The areas under the curve and their 95% confidence intervals for 5-year predictions for CBCRisk-Mastectomy and CBCRisk were 0.62 (0.59, 0.65) and 0.58 (0.55, 0.61), respectively.

**Conclusions:** CBCRisk-Mastectomy may aid clinicians in counseling BC patients scheduled for mastectomy, enabling improved decision-making regarding CPM.

## Introduction

Women affected with unilateral breast cancer (BC) are naturally worried about the risk of developing cancer in the opposite, unaffected breast, referred to as contralateral breast cancer (CBC). As a preventive measure, unilateral BC patients whose treatment involves mastectomy (surgical removal of the affected breast) may choose to undergo a contralateral prophylactic mastectomy (CPM), i.e., removal of the contralateral unaffected breast, at the same time as the mastectomy for the breast with the initial BC. Since the late 1990s, CPM rates have risen significantly in the United States (US), with a nearly six-fold increase between 1998 and 2011^1^. For US women aged 20-44 years, diagnosed with invasive unilateral early-stage BC that is treated with surgery, CPM rates rose from 10.5% in 2004 to 33.3% in 2012, reaching as high as 42% in some US states ^2^. A recent study conducted on 55,060 unilateral BC patients in the US reported a significant increase in CPM rates from 2016 to 2019, with the highest rates among younger women^3^. Another recent study found tripling of the CPM rate from 21.8% (in 2000–2009) to 64.4% (in 2017–2021) over two decades in a rural population^4^.

Although CPM is effective in reducing CBC risk, recent advances in adjuvant systemic therapies have also contributed to a decline in CBC incidence. In the US, CBC incidence decreased by 3% annually from 1985 to 2006^5^. A systematic review of nine studies, stratified by the median year of first BC diagnosis as 1989-1994, 1995-1999, and 2000-2003, reported declining median annual CBC incidence rates of 0.5%, 0.4%, and 0.2%, respectively^6^. The standardized incidence ratio of CBC among BC patients who did not undergo CPM compared to BC patients in the general population dropped from 2.49 in 1992-1997 to 1.57 in 2010-2015, suggesting that the advances in systemic therapies may be the primary driver of this decline^7^.

CPM is an aggressive approach for CBC prevention, yet it is not clear that it prolongs overall survival among BC patients without BRCA1/2 gene mutation^8-10^. Although several studies have reported improved overall survival among women with CPM, these are likely attributable to unmeasured confounders, and thus, are not causal^11^. Affluent non-Hispanic White women, those with private insurance, or higher socioeconomic status are more likely to opt for CPM followed by breast reconstruction^12-14^. While CPM may be medically justifiable for women at a high risk of CBC, there is no evidence of benefit for those with early-stage unilateral BC and low-to-average CBC risk^15^. The physical, psychological, and financial impacts of CPM can be substantial, including a lengthy recovery period, loss of breast sensation, and potential need for re-operations^16,17^. Compared to unilateral mastectomy, bilateral mastectomy carries higher risks of post-operative complications, more emergency room visits during the immediate post-operative period, and longer hospital stays^17-19^.

The decision to undergo CPM is often driven by a desire to reduce CBC risk, alleviate anxiety about CBC, and achieve breast symmetry. Yet risk reduction is minimal for most women because of their low initial CBC risk (notable exceptions are BRCA1/2 carriers or patients who have received mantle radiotherapy for Hodgkin lymphoma)^18,20,21^. Informing women about their risk of developing CBC based on personal characteristics can help them make more informed decisions about choosing the optimal surgical treatments, and, if CBC risk is low, avoid unnecessary CPMs^22^, and reduce anxiety of CBC^23,24^.

CBCRisk is an existing risk prediction model that estimates the absolute risk of developing CBC in women with unilateral BC^23,25^. It was trained on nationally representative data from the Breast Cancer Surveillance Consortium (BCSC) and the National Cancer Institute’s (NCI’s) Surveillance, Epidemiology, and End Results (SEER 17) database^26,27^. The model provides CBC risk predictions for all unilateral BC patients and was built using the same methodology as the NCI’s Breast Cancer Risk Assessment Tool (BCRAT), also known as “the Gail Model”, which predicts personalized risk of a first BC^28^. However, only those BC patients who undergo mastectomy for treatment of their initial BC, typically opt for CPM. Therefore, a CBC model trained solely on such patients is likely to be more useful for decision making about CPM. In this study, we developed CBCRisk-Mastectomy, which is specifically tailored for patients undergoing mastectomy and considering CPM.

## Data Description

### BCSC data

BCSC is a network of US breast imaging registries; this study used data from 5 active and 2 historical registries ^26^. Our study cohort included women who received their first BC diagnosis at ages 18-88 years between 1995 and 2009 and whose first BC was either DCIS or invasive. Upon applying the exclusion criteria described in the online supplement, the study cohort consisted of 24,962 women.

A woman was classified as a CBC case if she was diagnosed with invasive BC or DCIS in the opposite healthy breast at least six months after her initial BC diagnosis. Each CBC case was matched to three controls based on race, year of diagnosis, and equal or longer follow-up time compared to the case. Follow-up started at the time of initial BC diagnosis and ended at the time of CBC diagnosis for a case and at the time of censoring or death for controls. The final analytic dataset included 665 cases and 1995 controls, with 25 potential predictors (all categorical). Further preprocessing described in the supplement reduced the number of predictors to twenty. BRCA1/2 status or family history beyond first degree relatives is not available in the BCSC dataset. Additionally, while anti-estrogen therapy use is recorded, the specific type administered is unknown.

### SEER 17 data

We used the Incidence – SEER Research Plus Data (November 2023 submission, version 17), which provides population-level cancer information collected between 2000-2021 from 17 registries across the US^27^. As the month of diagnosis is not available in SEER, we classified a woman as a CBC case if she was diagnosed with BC in the opposite healthy breast at least one year after her initial BC diagnosis. Upon applying the exclusion criteria described in the online supplement, the final dataset included 445,266 women with BC of whom 15,984 developed CBC.

## Statistical Methods

We trained CBCRisk-Mastectomy on unilateral BC patients in BCSC and SEER who had undergone mastectomy as part of their treatment^26,27^. We refer to this subset of BC patients as “mastectomy patients”. The BCSC dataset has missing values for several important risk factors. Since excluding women with missing data would greatly reduce the sample size for fitting a multivariable regression model, in the training of CBCRisk, these women were included by allowing for an “unknown” category for most risk factors. However, as the unknown category is very heterogeneous, and breast cancer patients typically do know this information, here we imputed the missing data by employing multiple imputations as implemented in the Multiple Imputation by Chained Equations (MICE) package^29^. By restricting the training data to mastectomy patients and imputing missing data, we aim to provide more accurate CBC risk estimates to help BC patients make informed decisions about CPM. The observed data on predictors and CBC status were used to create ten imputed datasets, on which we fitted univariable and multivariable conditional logistic regression models.

### Building the relative risk (RR) model

To conduct an overall likelihood ratio test combining results from ten regression models with the same predictors fitted to the ten imputed datasets, we used a multiple imputation likelihood ratio test (MI-LRT)^30,31^ described in Supplement Section S3. For building the RR model, first we fitted univariate conditional logistic regression models for each of the twenty potential predictors, retaining those that had an MI-LRT p-value less than 0.25^23,24^. These predictors were then considered in constructing a multivariable (full) model. A risk factor was retained if the MI-LRT, comparing the full model with a reduced model (obtained by excluding that risk factor), yielded a p-value less than 0.10. Overall estimates of the log-odds ratios 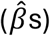 for association were obtained by averaging the corresponding estimates from the ten fitted models. These estimates approximate log-RRs as CBC a is rare event. Overall standard errors (SEs) were obtained by applying Rubin’s rule^32^. We computed 95% confidence intervals (CIs) for a regression coefficient as 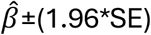. Estimates of RR and CIs were obtained by exponentiating the corresponding quantities.

### Building the absolute risk model

Following the statistical methods used in the Gail model^28^, CBCRisk^23^, and CBCRisk-Black^24^, we obtained age-specific composite incidence rates from SEER and attributable risks (AR) from the BCSC data. For the AR calculation, the study population was divided into thirteen age groups with 5-year intervals, specifically, < 30, 30–34, 35–39, …, 85-89. The incidence rates and ARs were then combined to estimate baseline age-specific hazard rates of CBC. We used SEER data from 2000-2021 to also estimate age-specific competing mortality rates from causes other than CBC. We then combined the RRs, ARs, age specific baseline CBC hazards, and competing mortality hazards as described in Gail mode^l28^ and the supplementary material of cbcrisk^23^ to obtain the absolute risk prediction model. Here, competing mortality events include death due to first BC as it precludes the possibility of CBC and all other deaths due to non-CBC causes. All such events of competing mortality are accounted for in the absolute risk calculation of CBC. A bootstrap method^33^ with 500 resamples was used to construct CIs for absolute risk estimates (see Supplement Section S3).

To evaluate the discriminatory performance of CBCRisk-Mastectomy, we calculated the area under the receiver operating characteristic curve (AUC) and its 95% CI for 5-year absolute risk predictions obtained using leave-one-group-out cross-validation (CV), as detailed in Supplement Section S3. A “group” refers to one case and its three matched controls. For comparison, we also computed the corresponding AUC for CBCRisk using the original (unimputed) matched case-control data, as the model allows an “unknown” category for missing predictor values.

The statistical software R (4.4.0) was used for all computations, including the MICE (3.18.0)^34^ and CBCRisk (2.0)^23^ packages. The package pROC (1.18.5) was used to estimate AUC and obtain its CI^35^. For conducting MI-LRT, we used the publicly available R code^30,36^.

## Results

Table 1 shows the distributions and coding of the twenty potential risk factors among cases and controls prior to imputation. The plausibility of the missing at random assumption underlying the analysis based on multiply imputed data was assessed and can be found in Supplement Section S4. Based on univariable analyses of the twenty risk factors, we retained twelve predictors: histologic type of first BC, lobular carcinoma in situ (LCIS), estrogen receptor (ER) status, breast density (obtained using the Breast Imaging Reporting and Data System (BI-RADS) method), age at first BC diagnosis, family history of BC (only first degree available in BCSC), age at first birth, body mass index (BMI), tumor stage, tumor size, anti-estrogen therapy, and radiation therapy. These twelve variables were used to fit the base model. To assess the significance of each variable in the presence of the other variables, we constructed twelve reduced models by excluding one variable at a time and applied MI-LRT. Tumor stage, tumor size, anti-estrogen therapy, and radiation therapy had p-values greater than 0.10 and hence were removed. The remaining eight variables were retained as important risk factors for predicting CBC at this stage.

**Table 1:** Distribution of potential risk factors associated with CBC among women diagnosed with a unilateral breast cancer who underwent a mastectomy. The variables associated with tumor characteristics and treatment are for the first BC.

| # | Risk Factors | Categories | Cases<br>(n = 665) | Controls<br>(n = 1995) |
| --- | --- | --- | --- | --- |
| **Personal Characteristics** |  |  |  |  |
| 1 | Age at first birth | 40+ | 8 (1.2%) | 8 (0.4%) |
|  |  | Nulliparous/<40 | 203 (30.5%) | 587 (29.4%) |
|  |  | Unknown | 454 (68.3%) | 1400 (70.2%) |
| 2 | Age at first BC diagnosis | <30 | 7 (1.1%) | 7 (0.4%) |
|  |  | 30-40 | 51 (7.7%) | 108 (5.4%) |
|  |  | 40+ | 607 (91.3%) | 1880 (94.2%) |
| 3 | Bilateral oophorectomy | Yes | 29 (4.4%) | 100 (5.0%) |
|  |  | No | 344 (51.7%) | 1021 (51.2%) |
|  |  | Unknown | 292 (43.9%) | 874 (43.8%) |
| 4 | Biopsy history | No | 275 (41.4%) | 843 (42.3%) |
|  |  | Yes | 133 (20.0%) | 343 (17.2%) |
|  |  | Unknown | 257 (38.6%) | 809 (40.6%) |
| 5 | BMI | Obese | 42 (6.3%) | 92 (4.6%) |
|  |  | Overweight | 47 (7.1%) | 128 (6.4%) |
|  |  | Normal/Underweight | 69 (10.4%) | 240 (12.0%) |
|  |  | Unknown | 507 (76.2%) | 1535 (76.9%) |
| 6 | Breast density | Extremely dense | 40 (6.0%) | 85 (4.3%) |
|  |  | Heterogeneously dense | 160 (24.1%) | 386 (19.3%) |
|  |  | Scattered | 111 (16.7%) | 358 (17.9%) |
|  |  | Almost entirely fat | 6 (0.9%) | 37 (1.9%) |
|  |  | Unknown | 348 (52.3%) | 1129 (56.6%) |
| 7 | Family history of BC | Yes | 79 (11.9%) | 169 (8.5%) |
|  |  | No | 236 (35.5%) | 761 (38.1%) |
|  |  | Unknown | 350 (52.6%) | 1065 (53.4%) |
| 8 | Hormonal use | No | 249 (37.4%) | 726 (36.4%) |
|  |  | Yes | 73 (11.0%) | 228 (11.4%) |
|  |  | Unknown | 343 (51.6%) | 1041 (52.2%) |
| 9 | Menopausal status | Post | 277 (41.7%) | 819 (41.1%) |
|  |  | Peri | 6 (0.9%) | 10 (0.5%) |
|  |  | Pre | 69 (10.4%) | 234 (11.7%) |
|  |  | Unknown | 313 (47.1%) | 932 (46.7%) |

| Tumor characteristics of first BC |  |  |  |  |
| --- | --- | --- | --- | --- |
| 10 | ER status | Negative | 142 (21.4%) | 338 (16.9%) |
|  |  | Positive | 327 (49.2%) | 1075 (53.9%) |
|  |  | Unknown | 196 (29.5%) | 582 (29.2%) |
| 11 | PR status | Positive | 297 (44.7%) | 928 (46.5%) |
|  |  | Negative | 173 (26.0%) | 477 (23.9%) |
|  |  | Unknown | 195 (29.3%) | 590 (29.6%) |
| 12 | HER2 status | Positive | 21 (3.2%) | 58 (2.9%) |
|  |  | Negative | 68 (10.2%) | 210 (10.5%) |
|  |  | Unknown | 576 (86.6%) | 1727 (86.6%) |
| 13 | First BC type | DCIS | 65 (9.8%) | 132 (6.6%) |
|  |  | Invasive | 600 (90.2%) | 1863 (93.4%) |
| 14 | High grade | No | 251 (37.7%) | 714 (35.8%) |
|  |  | Yes | 298 (44.8%) | 947 (47.5%) |
|  |  | Unknown | 116 (17.4%) | 334 (16.7%) |
| 15 | LCIS | Yes | 16 (2.4%) | 18 (0.9%) |
|  |  | No | 639 (96.1%) | 1966 (98.5%) |
|  |  | Unknown | 10 (1.5%) | 11 (0.6%) |
| 16 | Tumor stage | Advanced primary/Metastatic | 111 (16.7%) | 232 (11.6%) |
|  |  | Early | 489 (73.5%) | 1563 (78.3%) |
|  |  | Unknown | 65 (9.8%) | 200 (10.0%) |
| 17 | Tumor size | T1 | 250 (37.6%) | 825 (41.4%) |
|  |  | T0 | 5 (0.8%) | 27 (1.4%) |
|  |  | T2 | 174 (26.2%) | 580 (29.1%) |
|  |  | T3 | 56 (8.4%) | 118 (5.9%) |
|  |  | T4 | 16 (2.4%) | 33 (1.7%) |
|  |  | TIS | 121 (18.2%) | 294 (14.7%) |
|  |  | Unknown | 43 (6.5%) | 118 (5.9%) |
| Treatment of first BC |  |  |  |  |
| 18 | Anti-estrogen therapy | No | 474 (71.3%) | 1351 (67.7%) |
|  |  | Yes | 156 (23.5%) | 550 (27.6%) |
|  |  | Unknown | 35 (5.3%) | 94 (4.7%) |
| 19 | Chemotherapy | No | 390 (58.6%) | 1213 (60.8%) |
|  |  | Yes | 248 (37.3%) | 703 (35.2%) |
|  |  | Unknown | 27 (4.1%) | 79 (4.0%) |
| 20 | Radiation therapy | No | 521 (78.3%) | 1655 (83.0%) |
|  |  | Yes | 131 (19.7%) | 309 (15.5%) |
|  |  | Unknown | 13 (2.0%) | 31 (1.6%) |

Before finalizing the model, we explored other reduced models obtained by systematically excluding different subsets of the four non-significant variables and performing MI-LRTs to evaluate the significance of the excluded set of variables. This analysis led to the inclusion of tumor stage in the final model. Additionally, we examined interaction effects between pairs of risk factors for which sufficient data were available for each combination level. However, none of the interactions were significant.

Table 2 shows the odds ratios and 95% CIs for the nine risk factors in the final RR model. Having a DCIS diagnosis for the first BC type and the presence of LCIS were associated with a higher risk of developing CBC. CBC risk was also elevated in BC patients with negative ER status, family history of BC, higher BMI, denser breasts, younger age at first BC diagnosis, and older age at first birth. Furthermore, women whose first BC was diagnosed at an advanced/metastatic stage had an increased risk of developing CBC. We note that the confidence interval for the less than 30 years category of age at first diagnosis is very wide due to the small number of cases (*n* = 7) in this group. For model stability, we had to merge two consecutive categories for age at first birth, BMI, and stage as one of the categories had low sample size. Table S1 shows results without the merging, which are not substantially different.

**Table 2:** Risk factors associated with CBC risk in the final relative risk model among women diagnosed with a unilateral breast cancer who underwent a mastectomy.

| # | Risk Factors | Categories | Odds Ratio* | 95% CI* |
| --- | --- | --- | --- | --- |
| 1 | Age at first birth | 40+ | 2.07 | 1.19, 3.57 |
|  |  | Nulli/<40 | 1 |  |
| 2 | Age at first diagnosis | <30 | 3.07 | 0.94, 10.00 |
|  |  | 30-40 | 1.39 | 0.95, 2.04 |
|  |  | 40+ | 1 | - |
| 3 | BMI | Obese | 2.05 | 1.50, 2.79 |
|  |  | Overweight | 1.51 | 1.30, 2.20 |
|  |  | Normal/Underweight | 1 |  |
| 4 | Breast density | Extremely dense | 3.22 | 1.44, 7.24 |
|  |  | Heterogeneously dense | 2.97 | 1.35, 6.54 |
|  |  | Scattered | 1.96 | 0.86, 4.47 |
|  |  | Almost entirely fat | 1 |  |
| 5 | Family history of BC | Yes | 1.53 | 1.11, 2.10 |
|  |  | No | 1 |  |
| 6 | ER status | Negative | 1.35 | 1.03, 1.78 |
|  |  | Positive | 1 |  |
| 7 | First BC type | DCIS | 1.39 | 1.12, 1.74 |
|  |  | Invasive | 1 |  |
| 8 | LCIS | Yes | 2.51 | 1.22, 5.18 |
|  |  | No | 1 |  |
| 9 | Tumor stage | Advanced primary/Metastatic | 1.46 | 1.11, 1.91 |
|  |  | Early BC | 1 |  |

\*Odds ratios and 95% CIs are pooled estimates obtained using ten imputed datasets.

Among the nine risk factors in the final model, histologic type of first BC, ER, breast density, age at first diagnosis, family history of BC, and age at first birth are present in CBCRisk. Instead of LCIS, CBCRisk includes high-risk pre-neoplasia, which combined LCIS and atypical hyperplasia. First, BC type in CBCRisk has three categories, while CBCRisk-Mastectomy combines pure DCIS and invasive DCIS into a single category, DCIS, reducing the total to two categories. Additionally, anti-estrogen therapy use is included in CBCRisk, but not in CBCRisk-Mastectomy.

Figure 1 presents the SEER age-specific composite CBC incidence and competing mortality rates (per 1,000 person-years). The CBC incidence rates ranged from 2.58 to 4.89, with the lowest rate observed in the [18, 30) age group and the highest in the [75, 80) age group. The rate generally increased up to age 80 and then declined. Non-CBC mortality rates ranged from 16.42 to 107.90 per 100,000, with the lowest rate in the [45, 50) age interval and the highest in the [85, 90) interval.

**Figure 1:**
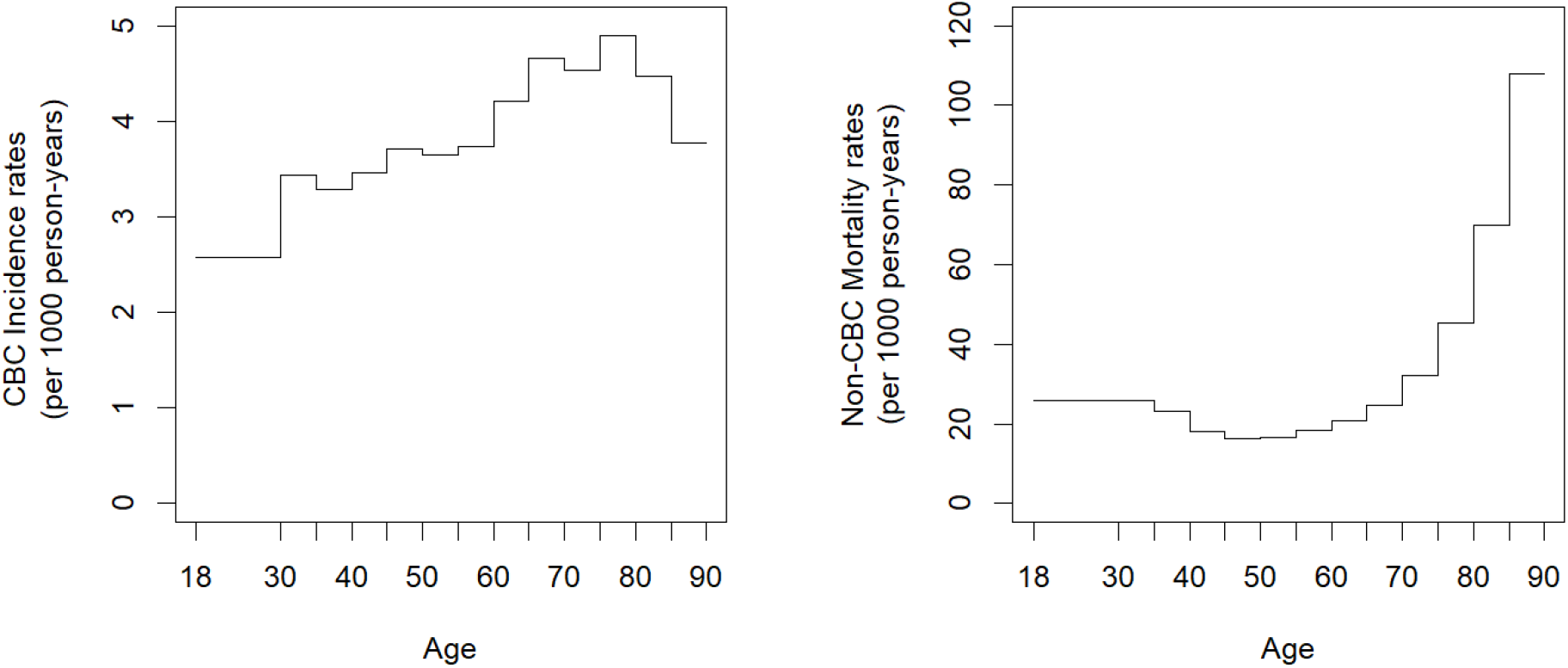
Age-specific composite incidence rates of CBC (left) and non-CBC mortality rates (right) estimated using SEER data. Age (in years) on the horizontal axis can be interpreted as age at counseling.

The average AUC (with pooled 95%CI) for 5-year absolute risk predictions from CBCRisk-Mastectomy across the ten imputations computed using Rubin’s rule was 0.620 (0.586, 0.654). In comparison, for CBCRisk, the AUC computed by applying the standard asymptotic formula to the original data was 0.577 (0.547, 0.608). This shows that CBCRisk-Mastectomy has better discrimination ability than CBCRisk for the target population of mastectomy patients. A statistical test for difference of AUCs from the two models is not straightforward to calculate due to the difference in datasets and methods. Further, calibration measures such as the ratio of expected to observed numbers of cases cannot be meaningfully computed for matched case-control data, as the observed proportion of cases is fixed by design, which is 1/4 for our data.

Figure 2 shows the distributions of 5-year absolute risk predictions separately among cases and controls for both models. CBCRisk-Mastectomy better discriminates cases from controls as reflected in a larger separation in their risk distributions. This larger difference leads to the higher AUC of CBCRisk-Mastectomy.

**Figure 2:**
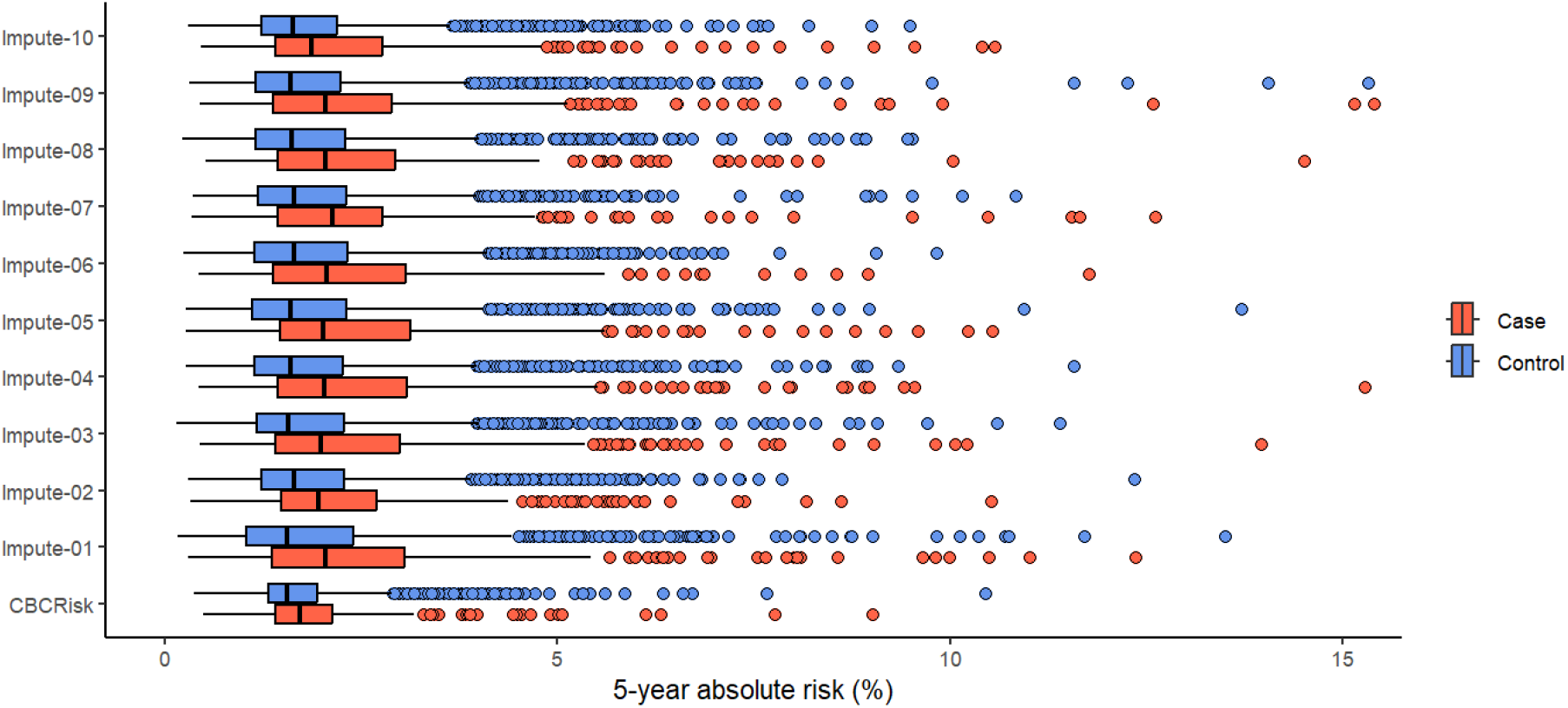
Leave-one-group-out 5-year absolute risk predictions using CBCRisk-Mastectomy based on ten imputed datasets and CBCRisk. Here absolute risk values above 15% absolute risk are excluded to ensure a smaller scale and better readability. The full figure without this restriction is given in the online supplement as Figure S1.

In Table 3, we present 5-year absolute risk predictions of CBC, using both CBCRisk-Mastectomy and CBCRisk models, for eight women selected from those with complete risk factor profile in BCSC data, i.e., those without any missing data on the predictors. These examples cover a broad spectrum of risk factor combinations and current ages. The table clearly illustrates that CBC risk varies substantially based on a woman’s specific risk profile. The first row represents a woman with each risk factor at the most common category, which is also the reference category for all risk factors in CBCRisk-Mastectomy except breast density. In this case, the 5-year CBC risk is 1.06%, indicating that the risk for a typical BC patient is generally low. The other examples further show that CBC risk remains low across different risk factor combinations. Only in the last example the risk exceeds 10% and there is, in fact, only one woman with that risk profile in the subset of women with complete information on all nine risk factors. Additionally, for the last example, the 5-year absolute risk of CBC predicted using CBCRisk-Mastectomy is notably higher than that by CBCRisk. This elevated risk may be attributed to differences in the risk factor profiles incorporated in the models. In particular, while CBCRisk considers anti-estrogen therapy as a risk factor, it does not include BMI or tumor stage, both of which are included in CBCRisk-Mastectomy.

**Table 3:**
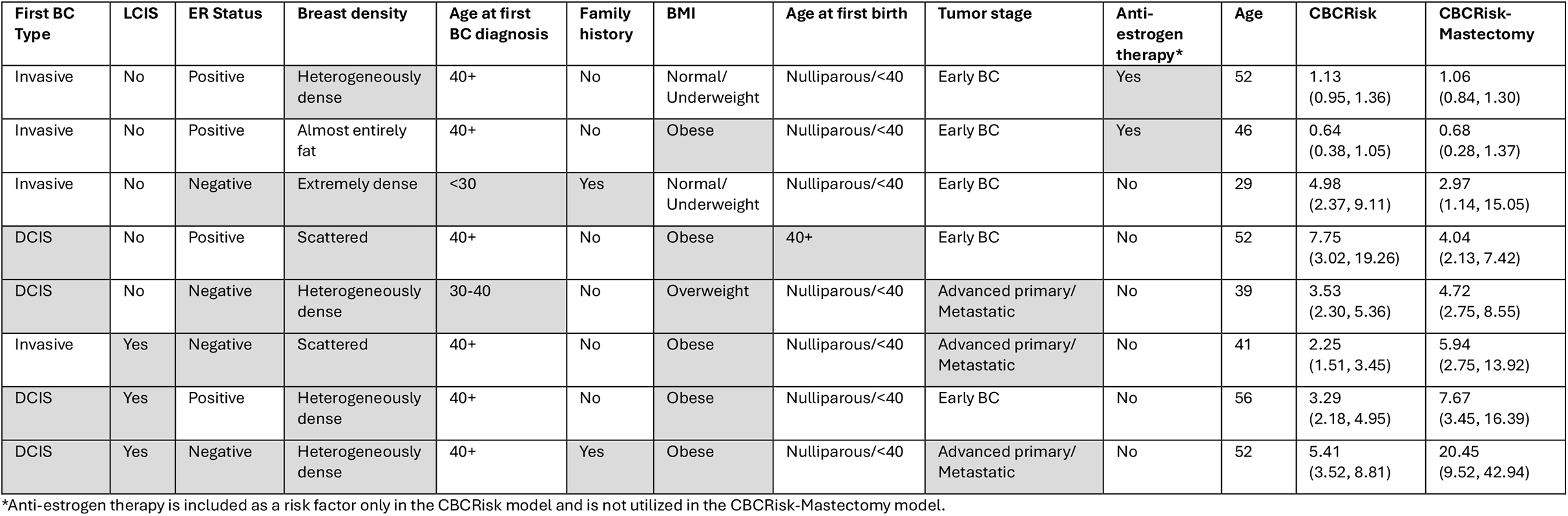
Examples of 5-year CBC risk prediction with 95% confidence intervals in parenthesis using CBCRisk-Mastectomy and CBCRisk. The highlighted cells represent non-baseline categories.

## Discussion

BC patients scheduled to undergo mastectomy typically decide, in consultation with their surgeon, whether to remove their other, unaffected breast as well (i.e., whether to undergo CPM) at the same time as the surgery for the original BC. To assist in making this difficult decision we have developed CBCRisk-Mastectomy, a personalized CBC risk prediction model specifically tailored to such patients. By utilizing data from two nationally representative datasets — BCSC and SEER — our model is applicable to the general US population of unilateral BC female patients (except those with genetic mutations, strong family history, or prior mantle radiation). CBCRisk-Mastectomy incorporates nine predictors: first BC type (DCIS or invasive), presence of LCIS, ER status, breast density, age at first diagnosis, family history of BC, age at first birth, BMI, and tumor stage, all well established CBC risk factors. For instance, DCIS, LCIS, and advanced-stage initial BC are associated with a higher risk of developing CBC^37,38^. Previous studies have also shown that negative ER status, having a family history of BC, denser breasts, and higher BMI are associated with increased CBC risk^39-41^, as are younger age at first BC diagnosis and older age at first childbirth^42^.

It is important to note that CBCRisk-Mastectomy is specifically intended for patients who are scheduled to undergo mastectomy and are considering CPM at the same time to help them in making this decision. For women undergoing lumpectomy, CBCRisk can be used as it predicts the risk of CBC for any women with unilateral BC. It is difficult for us to provide thresholds for identifying women at high risk for CBC. In this regard, we note that Gail and Jatoi (2022) classified women with 10-year CBC risk of 0-2.49% risk as low risk, with 2.5-9.9% risk as medium risk, and with 10% risk or higher as high risk^43^.

As anti-estrogen therapy is known to lower CBC risk but was not selected as an important predictor during our model building steps, we explored the possibility of adding it back to the final model.

However, inclusion of anti-estrogen therapy did not improve prediction accuracy (MI-LRT p-value = 0.28), likely because its effect was largely captured by ER status and first BC type. These two variables are highly correlated with anti-estrogen therapy (p-values ≈ 0) and were stronger predictors of CBC risk than anti-estrogen therapy. Thus, they got included in the final CBCRisk-Mastectomy model. For model training, we had also tried MI-LASSO technique, but it retained all the variables, which is a known limitation of the method and thus was not pursued further^44^. We also tried fitting the model on original data. However, there are only 61 cases with complete information on all nine risk factors of CBCRisk-Mastectomy with five or fewer observations for several categories of multiple risk factors, which prevented model fitting to the complete-case data.

Comparing CBCRisk-Mastectomy with CBCRisk demonstrated improved discrimination of the new model for mastectomy patients, as indicated by the higher AUC. Seven out of the eight risk factors in the CBCRisk model are also in the CBCRisk-Mastectomy model, with the only exception being anti-estrogen therapy. CBCRisk-Mastectomy also incorporates BMI and tumor stage. Additionally, while CBCRisk includes an “unknown” category for most predictors ^23^, CBCRisk-Mastectomy does not have “unknown” predictor categories, as we imputed the missing data before training the model. Even though having an “unknown” category allows a model to be applicable to patients with missing risk factors, it can be detrimental to model training because missing categories are inherently mixtures of other categories and thus highly heterogeneous. By using multiple imputation before training, we reduced this heterogeneity, which as our results show, increased model accuracy.

It is noteworthy that the absolute risk of CBC is generally low for most women, even among those with multiple risk factors present, as shown in Table 3. Moreover, risks of regional and distant recurrences of BC should also be taken into account in CPM decision-making, given that CPM does not reduce these risks. Although CBC is the most common second malignant tumor among women diagnosed with BC, it occurs less frequently than recurrence of the initial BC^45^. Younger women face a higher lifetime risk of CBC, but this risk decreases with age, while the risks of regional and distant recurrence, as well as death from other causes, increase^46^. Gail and Jatoi (2022) highlighted the importance of evaluating all these risks when counseling BC patients on the risks and benefits of CPM^43^. They emphasize that a comprehensive approach that considers personalized risks of CBC, regional recurrence, distant recurrence, and non-CBC mortality should guide the decision-making process regarding CPM^15^.

Even though the AUC of 0.62 for CBCRisk-Mastectomy reflects moderate discriminatory ability, it is similar to that of other established models that predict breast cancer incidence in high risk women or the general population. For example, CBCRisk reported AUCs of 0.62 and 0.61 in external validations^25^ and the widely used Gail model for predicting BC risk for the general US female population had a pooled AUC of 0.60 (95% CI: 0.58–0.62)^47^. Similarly, PredictCBC-2.0A, designed for high-risk BC patients with BRCA1/2 mutation, and PredictCBC-2.0B, designed for the general population of BC patients, had AUCs of 0.66 and 0.59, respectively^48^. Despite moderate discriminatory performance, these models are currently in use in clinical settings, supporting the potential utility of CBCRisk-Mastectomy for individualized CBC risk assessment and surgical decision-making.

Despite the benefits of the CBCRisk-Mastectomy model, some limitations should be acknowledged. First, the high percentage of missing data for certain variables necessitated using imputations, which induce some degree of uncertainty. A second limitation is the absence of BRCA1/2 mutation information in the BCSC data and hence in the model. Even though younger age at first BC diagnosis combined with a family history of BC may partly capture the effect of germline pathogenic variants, adding genetic information directly in the model may improve model accuracy as reflected in the fact that PredictCBC 2.0A has much higher AUC than that of PredictCBC 2.0B (as mentioned above). An important future work will be to update the model with BRCA1/2 status once data with this information become available.

Another limitation is that the majority of our study population consisted of White non-Hispanic US women, which may limit the generalizability of the model to other races and ethnicities. As we matched CBC cases and controls on race, race cannot be a predictor in the model. Earlier we had developed CBCRisk-Black (based on 186 cases from BCSC), which can be used for Black BC patients^24^. In mastectomy patients, only 82 cases are Black; such small sample sizes make it challenging to building race-specific models. As the month of diagnosis is not available in the SEER data, we defined CBC as a malignancy occurring in the contralateral breast at least one year after the initial BC diagnosis, while for BCSC, we used a 6 months duration after the initial breast cancer – using 12 months duration would have led to loss of 79 cases in BCSC. An important limitation is that due to the matched case-control design of our study, we could not assess calibration, i.e. bias of the model predictions, which is typically done by comparing the expected number of cases produced by the model with the observed number of cases in a prosepctive cohort. Finally, the BCSC data used in this study were collected from women diagnosed between 1995 to 2009. Therefore, validating the model, including assessing its calibration on newer external data is warranted. CBCRisk-Mastectomy will be ready for clinical implementation after an external validation of the model.

In conclusion, CBCRisk-Mastectomy is a potentially valuable tool to assist clinicians and patients in making informed decisions regarding CPM and tailoring treatment and preventive choices by utilizing personalized risk estimates. Given the physical, psychological, and financial burdens associated with CPM, providing an evidence-based tool for personalized risk prediction is crucial for optimizing care and minimizing unnecessary CPM. Future research should focus on validating this model on external or more recent data and exploring its integration into clinical practice.

## Acknowledgements

Data collection was supported in part by the National Cancer Institute-funded Breast Cancer Surveillance Consortium (P01CA154292). The collection of BCSC cancer and vital status data was supported in part by several state public health departments and cancer registries (https://www.bcsc-research.org/work/acknowledgement). All statements in this report, including its findings and conclusions, are solely those of the authors and do not necessarily represent the views of the National Cancer Institute or National Institutes of Health. We thank the participating women, mammography facilities, and radiologists for the data they have provided.

You can learn more about the BCSC at: http://www.bcsc-research.org/. The authors are thankful to Dr. Debu Tripathy for insightful discussion on this topic. The authors also thank two reviewers for their thorough and thoughtful review of the manuscript. Addressing their comments has led to an improved article.

## Funding

This project was partially funded by the National Cancer Institute at the National Institutes of Health (Grant Number R21CA186086-01).

## Data availability

Instructions on how to request BCSC data can be found at https://www.bcsc-research.org/. SEER data can be requested at https://seer.cancer.gov/.

## Software

CBCRisk-Mastectomy is implemented in R package CBCRisk version 3.0 available at personal.utdallas.edu/~swati.biswas/ and github.com/ihsajal/CBCRisk.

## Conflict of Interest

The authors report no proprietary or commercial interest in any product mentioned or concept discussed in this article.

## Notes

### Competing Interest Statement

The authors have declared no competing interest.

### Author Declarations

IRB of the Office of Research and Innovation at the University of Texas at Dallas gave the ethical approval for this work.

