## Supplemental_Files for "CBCRisk-Mastectomy: A Risk Prediction Tool to Aid Contralateral Prophylactic Mastectomy Decision Making"

**by**

**Ibrahim Hossain Sajal, Ruth M. Pfeiffer, Ismail Jatoi, Mitchell H. Gail, Reena S. Cecchini, Pankaj K. Choudhary, and Swati Biswas**

### S1. Additional Details for BCSC Data

**Exclusion Criteria:** We excluded women satisfying any of the following criteria: (a) first BC was bilateral or had unknown laterality, (b) unknown laterality at a second or a later BC diagnosis if all previous diagnoses reported the same laterality, (c) lacking histological confirmation of malignancy for the first BC diagnosis, (d) radiation therapy before the first BC diagnosis, (e) first BC type was not DCIS or invasive, and (f) unavailable or ill-defined race. Further, we restricted the dataset to women who had undergone mastectomy for treatment of their first BC.

**Preprocessing:** Of the 25 potential predictors, five predictors were excluded: four (personal history of ovarian cancer, first-degree family history of ovarian cancer, family history of ovarian cancer other than first degree, and personal history of atypical hyperplasia) due to over 93% missing data, and one (number of positive lymph nodes) because it had only three cases in the “1+” category. For some predictors, if there were insufficient numbers of cases in certain categories, we combined adjacent categories.

### S2. Additional Details for SEER Data

**Exclusion Criteria:** We applied the same exclusion criteria as those for the BCSC cohort, except for prior radiation therapy, as this information is not available in SEER.

### S3. Additional Details of Statistical Methods

**MI-LRT implementation**: For each imputed dataset, we computed the complete-data likelihood ratio statistic comparing the null model to an alternative model. These likelihood ratio statistics and associated estimates are then pooled across the multiple imputations using combining rules that account for both within-imputation and between-imputation variability[1,2]. The pooled test statistic is adjusted so that it follows (approximately) an F distribution (or a chi-square in large samples), allowing us to assess significance while properly incorporating uncertainty due to missing data.

**Confidence interval for absolute risk:** We applied a bootstrap method to calculate CI for the absolute risk of CBC. Following the MI-Boot (pooled sample) methodology outlined by[3], we generated 500 bootstrap resamples for each of the ten imputed datasets, estimated RRs and ARs for each bootstrap resample, and predicted 5-year absolute risk. In this way, we obtained 5,000 absolute risk estimates of CBC. Percentiles of the empirical bootstrap distribution function were used to compute CIs.

**Leave-one-group-out-cross validation**: Each case was grouped together with its three matched controls, resulting in 665 groups comprising a total of 665 cases and 1995 corresponding controls. This method involved fitting the RR model with the final set of predictors using all groups except one, which was set aside as a test group. The regression coefficients from this fitted model and the corresponding estimates of the baseline hazard were then used to predict the absolute risks for the women in the test group. This procedure was repeated by sequentially setting aside a different group each time. Thus, at the end, the absolute risk estimates for all women in the dataset were available, which were then used to compute the AUC. This CV method was applied separately to each of the ten imputed datasets, resulting in ten estimates of AUC and its 95% CIs. For each AUC estimate, the SE was derived from the corresponding CI assuming normality. An overall 95% CI for AUC was calculated by pooling the ten SEs using Rubin's rule[4].

### S4. Evaluation of Missing at Random (MAR) Assumption

We follow two suggestions in the mice package[5] to examine the plausibility of the MAR assumption that justifies using multiple imputation. First, we compare the observed and the imputed data distributions of each of the seven variables in the model that have missing data. The results presented in Figure S2 show that the two distributions look quite similar for all variables.

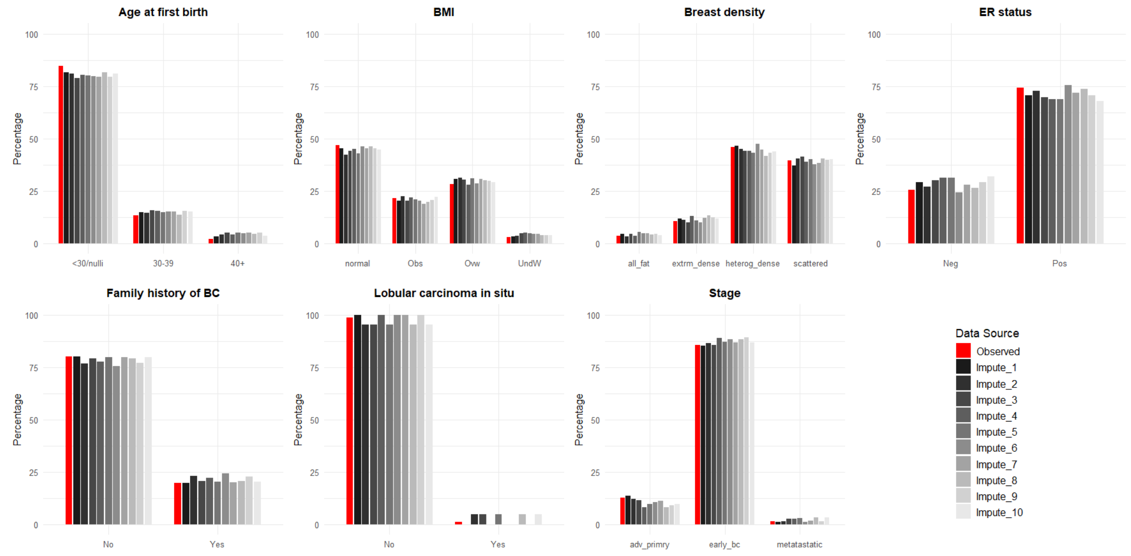

*Figure S1: Distributions of seven risk factors across observed and ten imputed datasets.*

Next, we compare the two distributions conditional on the propensity score quartiles. Figure S3 shows the results for breast density, which has around 55% missing values. We do not see substantial differences in the distributions in observed and imputed data. Similar conclusions hold for the other variables as well (figures not shown). Altogether, these observations point to the plausibility of the MAR assumption.

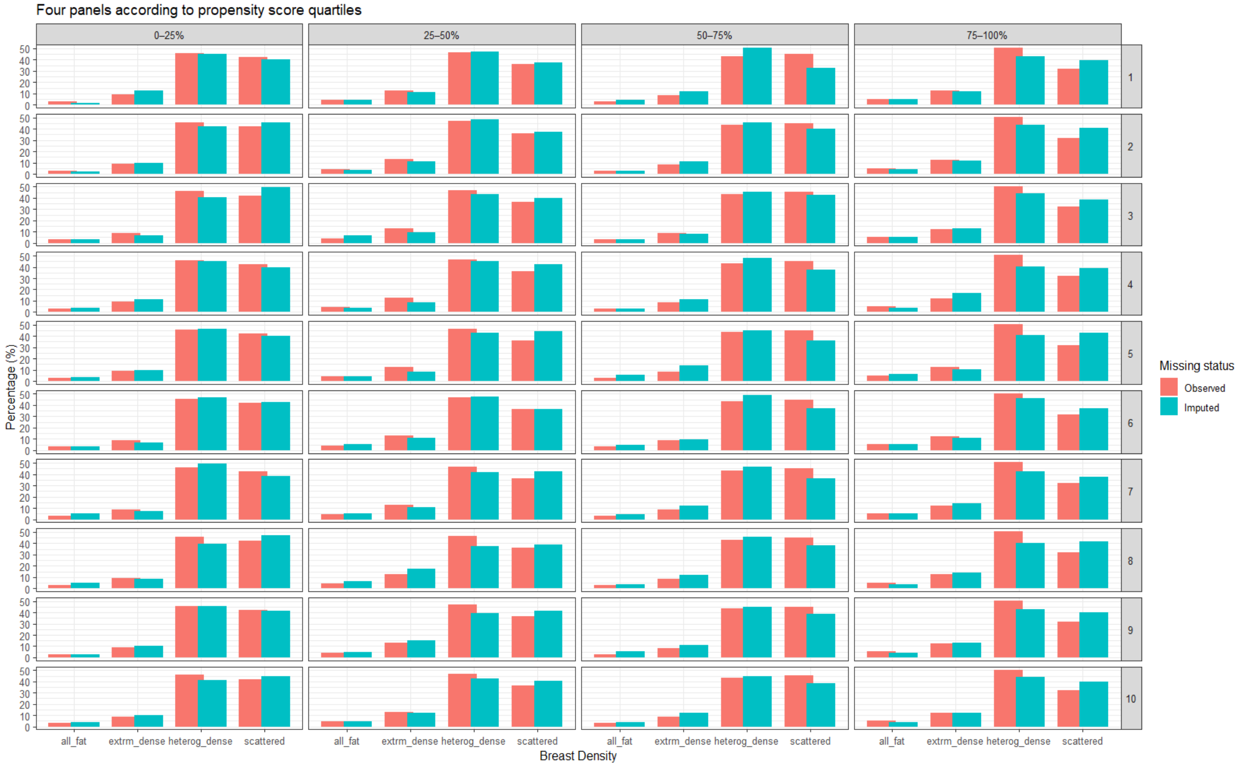

*Figure S2: Distribution of Breast Density at propensity score quartiles across observed and imputed data. The four vertical panels refer to the four quartiles of the propensity score, whereas the ten horizontal panels refer to the distributions from ten imputed datasets. The variable categories are: "all_fat" (almost entirely fat), "extrm_dense" (extremely dense), "heterog_dense" (heterogeneously dense), and "scattered".*

### S5. CBCRisk-Mastectomy Model: Inputs and Usage

The **cbcrisk** R package estimates age-specific absolute risk of contralateral breast cancer (CBC) among women with a prior breast cancer diagnosis. This study used the **CBCRisk-Mastectomy** model, which applies to women of any race who are scheduled to undergo mastectomy and are considering contralateral prophylactic mastectomy (CPM).

#### Function call

```
cbcrisk(mastectomy, race, profile, start.age, pred.year, print.output = TRUE)
```

#### Required inputs

- **mastectomy**: 1 (scheduled to undergo mastectomy; selects CBCRisk-Mastectomy)
- **race**: non-Hispanic Black (1) or otherwise (0); does not affect model choice when mastectomy = 1
- **profile**: numeric vector of length 9 containing category codes for the following risk factors (unknown values not allowed):
    1. Age at first birth: <40/nulliparous (1), ≥40 (2)
    2. Age at first breast cancer diagnosis: <30 (1), 30–39 (2), ≥40 (3)
    3. Body mass index: normal/underweight (1), overweight (2), obese (3)
    4. Breast density: extremely dense (1), heterogeneously dense (2), scattered (3), almost entirely fat (4)
    5. First-degree family history of breast cancer: yes (1), no (2)
    6. Estrogen receptor status: negative (1), positive (2)
    7. First breast cancer type: DCIS (1), invasive (2)
    8. Lobular carcinoma in situ (LCIS): yes (1), no (2)
    9. Tumor stage: early breast cancer (1), advanced primary/metastatic (2)
- **start.age**: age at counseling (18–89 years; must be ≥ age at first diagnosis)
- **pred.year**: prediction interval in years (e.g., 2, 5, 10), with predictions generated from start.age up to age 89
- **print.output**: logical indicator to print results to the R console (default = TRUE)

**Example**

```
cbcrisk(
  mastectomy = 1,
  race = 0,
  profile = c(1, 3, 2, 2, 1, 2, 2, 2, 1),
  start.age = 52,
  pred.year = 5
)
```

Table S1: Risk factor distributions among CBC cases and controls and log-odds before and after merging categories for BMI, Age at first birth, and Stage.

| | | | | Before Merging | | After Merging | |
|---|---|---|---|---|---|---|---|
| **Risk Factor** | **Category** | **Cases** | **Controls** | **log-odds** | **SE** | **log-odds** | **SE** |
| First BC type | DCIS | 65 (9.8%) | 132 (6.6%) | 0.3288 | 0.1137 | 0.3323 | 0.1135 |
| | Invasive | 600 (90.2%) | 1863 (93.4%) | Ref | | | |
| LCIS | Yes | 16 (2.4%) | 18 (0.9%) | 0.9519 | 0.3741 | 0.9217 | 0.3689 |
| | No | 639 (96.1%) | 1966 (98.5%) | Ref | | | |
| ER | Neg | 142 (21.4%) | 338 (16.9%) | 0.3037 | 0.141 | 0.3011 | 0.1401 |
| | Pos | 327 (49.2%) | 1075 (53.9%) | Ref | | | |
| Breast density | Extremely dense | 40 (6.0%) | 85 (4.3%) | 1.1551 | 0.421 | 1.1705 | 0.4129 |
| | Heterogeneously dense | 160 (24.1%) | 386 (19.3%) | 1.0937 | 0.4086 | 1.0881 | 0.4028 |
| | Scattered | 111 (16.7%) | 358 (17.9%) | 0.6724 | 0.4292 | 0.6713 | 0.4218 |
| | Almost entirely fat | 6 (0.9%) | 37 (1.9%) | Ref | | | |
| Age at first diagnosis | <30 | 7 (1.1%) | 7 (0.4%) | 1.1502 | 0.6092 | 1.1214 | 0.6028 |
| | 30-40 | 51 (7.7%) | 108 (5.4%) | 0.337 | 0.197 | 0.3303 | 0.1961 |
| | 40+ | 607 (91.3%) | 1880 (94.2%) | Ref | | | |
| Family history | Yes | 79 (11.9%) | 169 (8.5%) | 0.4273 | 0.1648 | 0.4231 | 0.1618 |
| | No | 236 (35.5%) | 761 (38.1%) | Ref | | | |
| BMI | Obese | 42 (6.3%) | 92 (4.6%) | 0.748 | 0.1612 | 0.7162 | 0.1584 |
| | Overweight | 47 (7.1%) | 128 (6.4%) | 0.4451 | 0.2049 | 0.4098 | 0.1927 |
| | Underweight | 5 (0.8%) | 14 (0.7%) | 0.407 | 0.4543 | Ref | Ref |
| | Normal | 64 (9.6%) | 226 (11.3%) | Ref | | | |
| Age at first birth | 40+ | 8 (1.2%) | 8 (0.4%) | 0.731 | 0.2843 | 0.725 | 0.2796 |
| | 30-39 | 30 (4.5%) | 78 (3.9%) | -0.0357 | 0.1779 | Ref | Ref |
| | <30/Nulliparous | 173 (26.0%) | 509 (25.5%) | Ref | | | |
| Stage | Advance primary | 101 (15.2%) | 207 (10.4%) | 0.4094 | 0.1479 | 0.3779 | 0.1377 |
| | Metastatic | 10 (1.5%) | 25 (1.3%) | 0.2379 | 0.4049 | | |
| | Early | 489 (73.5%) | 1563 (78.3%) | Ref | | | |

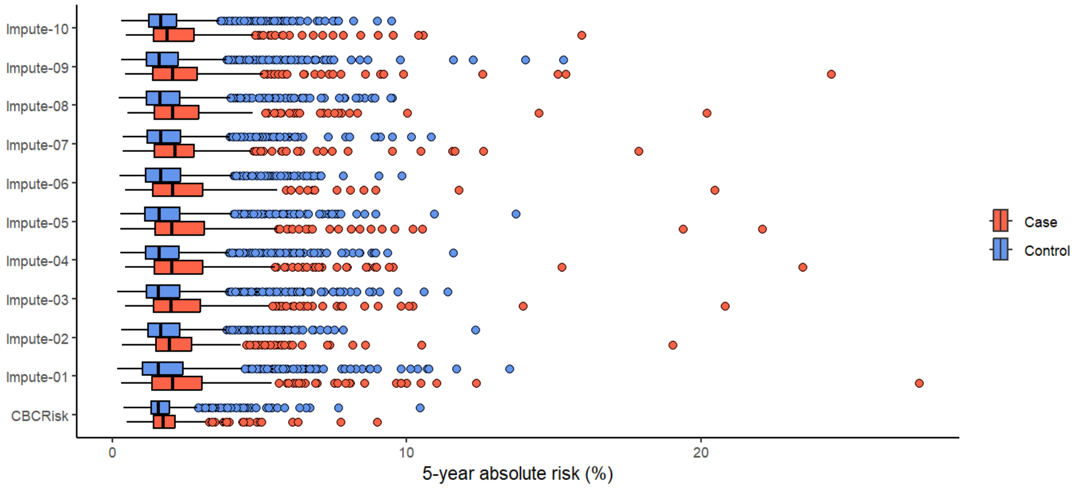

*Figure S1: Leave-one-group-out 5-year absolute risk predictions using CBCRisk-Mastectomy based on ten imputed datasets and CBCRisk.*
